# Reconstruction of SARS-CoV-2 outbreaks in a primary school using epidemiological and genomic data

**DOI:** 10.1101/2022.10.17.22281175

**Authors:** Cécile Kremer, Andrea Torneri, Pieter J. K. Libin, Cécile Meex, Marie-Pierre Hayette, Sébastien Bontems, Keith Durkin, Maria Artesi, Vincent Bours, Philippe Lemey, Gilles Darcis, Niel Hens, Christelle Meuris

## Abstract

Mathematical modeling studies have shown that repetitive screening can be used to mitigate SARS-CoV-2 transmission in primary schools while keeping schools open. However, not much is known about how transmission progresses within schools and whether there is a risk of importation to households. In this study, we reconstructed outbreaks observed during a prospective study in a primary school and associated households in Liège (Belgium) during the academic year 2020-2021. In addition we performed a simulation study to investigate how the accuracy of estimated weekly positivity rates in a school depends on the proportion of a school that is sampled in a repetitive screening strategy. We found that transmission occurred mainly within the school environment and that observed positivity rates are a good approximation to the true positivity rate, especially in children. This study shows that it is worthwile to implement repetitive testing in school settings, which in addition to reducing infections can lead to a better understanding of the extent of transmission in schools during a pandemic and importation risk at the community level.

## Introduction

During the early stages of the COVID-19 pandemic in the first half of 2020, several studies have investigated the contribution of children to SARS-CoV-2 transmission with results suggesting that schools did not play a substantial role in driving community transmission [1–3]. Mensah et al. [2] found that SARS-CoV-2 infections in school-aged children followed the same trend as adult cases and only declined after a national lockdown was implemented while keeping schools open, suggesting community transmission impacts transmission in schools. However, this study was based on data from routine symptom-based surveillance not fully capturing asymptomatic infections in children. Another study reported a positive correlation between community cases and cases in schools in England, with weak evidence suggesting that cases in schools lag behind community cases [3]. In contrast, a study in the US found that in-person schooling poses an increased risk of infection to household members, which can be reduced by implementing mitigation measures in schools [4]. Similar findings have been reported in Sweden [5]. In addition, the role of children has likely changed throughout the pandemic in light of the different variants of concern (VoC) circulating. In Belgium, the 2021-2022 academic year in primary schools started in September 2021 without any non-pharmaceutical intervention measures in place. When the school year progressed the number of SARS-CoV-2 cases rose and these interventions (i.e., mask wearing for teachers, isolation of symptomatic children, class closure after two cases) were reinstated. The rise of the Omicron variant in early 2022 caused a surge in case numbers, resulting in numerous school closures throughout the country.

In addition to structural interventions such as improving air quality in schools through better ventilation systems [6, 7], screening protocols such as repetitive and reactive screening can also aid in preventing the occurrence of large school outbreaks and consequent closures. In reactive screening, a classroom is tested and quarantined after a certain number of cases is observed, whereas in repetitive screening the entire classroom is tested regularly, regardless of whether a case is observed or not. A recent study using an agent-based model found that in a primary school population, weekly screening of 75% of unvaccinated children could reduce the number of cases by 34% on average, compared to symptom-based testing alone. In addition, the number of school days lost would be reduced up to 80% compared to reactive screening policies [8]. In line with this, another modeling study found that repetitive screening greatly reduced the attack rate in school, while reactive screening performed only slightly better than symptomatic isolation [9]. Furthermore, using a repetitive screening strategy allowed for increasing the class closure threshold without much impact on the attack rate. Besides limiting transmission, data obtained after implementing a repetitive screening strategy can be used to gain a better understanding of the extent of transmission within schools and the risk of children, as well as school employees, bringing infection into their households [10]. Such insights are crucial to ensure effective mitigation measures [6].

During the academic year 2020-2021, a prospective surveillance study using repetitive screening was set up in a primary school located in Liège (Belgium) with the objective to elucidate a better understanding of the role of children in SARS-CoV-2 transmission. Results from the first part of this study (September to December 2020) indicated transmission occurred mainly between children and among teachers within the school, with occasional spillover to their households [10]. Sequence data of rapidly evolving RNA viruses can provide valuable information on transmission events [11], as also illustrated specifically for SARS-CoV-2 [12]. In this study we make use of the whole genome sequences that were collected from September 2020 to June 2021 to aid in reconstructing the school outbreaks. Furthermore, using a simulation model, we investigate how the accuracy of estimated weekly positivity rates in a school depends on the proportion of a school that is sampled in a repetitive screening strategy.

## Results

### SARS-CoV-2 positivity in the study population

From September to December 2020, weekly screening for SARS-CoV-2 infection was performed through throat washing. Screening was increased to twice a week from January to June 2021. A description of the study design is provided in Methods. The total study population included 240 individuals, of which 88 children (36.7%) aged 5 to 13 years, 110 parents of these children (45.8%), 14 school employees (5.8%), 22 teachers (9.2%), and 5 participants (2.1%) that were both teacher and parent of a child included in the study. The children and parents made up 66 households, including 22 sibling pairs. The children and teachers were part of 20 class groups at the primary school and kindergarten level, and made up approximately 25% of the school population. Of the adults, 55.9% had received at least one vaccination dose by the end of the study, with 28.3% being fully vaccinated (Supplementary Fig. S1). Quantitative reverse transcription-PCR was used to detect SARS-CoV-2 and genome sequencing of positive samples was performed (see Methods). Over the entire study period, 61 individuals (25.4%) tested positive at least once for SARS-CoV-2, of which 22 children (25.0% of all participating children) and 39 adults (25.7% of all participating adults). There were two reinfections during the study period, concerning two adults. When accounting for clustering of individuals in classrooms and households using a mixed effects logistic regression model (see Methods), there was no significant difference in positivity for children compared to adults (conditional odds ratio 0.99 (bootstrap 95% CI 0.44 - 2.10), *p* = .979). The intraclass correlation (ICC) was 0.39 for household and 0.05 for classroom. Before January 2021, the overall positivity rate was 4.2% in adults (52/1249 samples) compared to 2.7% in children (18/667 samples). Between January and June 2021, the overall positivity rate was 0.6% in adults (25/4118 samples) compared to 1.7% in children (41/2465 samples). Overall, the positivity rate was 2.5% (218/8863 samples; Supplementary Fig. S2). Data on symptoms experienced by infected individuals were available for 59 cases. Over the full study period, 45 cases were symptomatic, while 14 remained asymptomatic, and children were not more often asymptomatic compared to adults (35.0% vs 16.2%, *p* = .184). Cycle threshold (Ct) values at diagnosis did not differ between children and adults in either time period, nor between time periods (*p* = 1.000 for week 1-15, *p* = .299 for week 16-40, *p* = .655 between periods; Supplementary Fig. S3). Ct values at diagnosis also did not differ between symptomatic and asymptomatic individuals (*p* = .592).

### Compliance to the study protocol

Among all participants, the median duration of participation was 38 weeks (IQR 25 - 40). Between September and December 2020, the median number of samples per participant was 12 (IQR 10 - 13), reflecting high adherence to the study protocol of one sample every week for 15 weeks. Adherence was in general a bit lower between January and June 2021 (week 16 to 40), with a median of 34 (IQR 21 - 39) samples per participant. Overall, adherence was higher among children and parents compared to school staff (Figure 1). Of the 235 participants included from January 2021 onward, 88 (37.5%) dropped out of the study before the last week of sampling. Among those who did not complete the study, 18.3% had previously tested positive for SARS-CoV-2 during the study period and 17.2% had received a first vaccination dose before their last sample.

**Figure 1:**
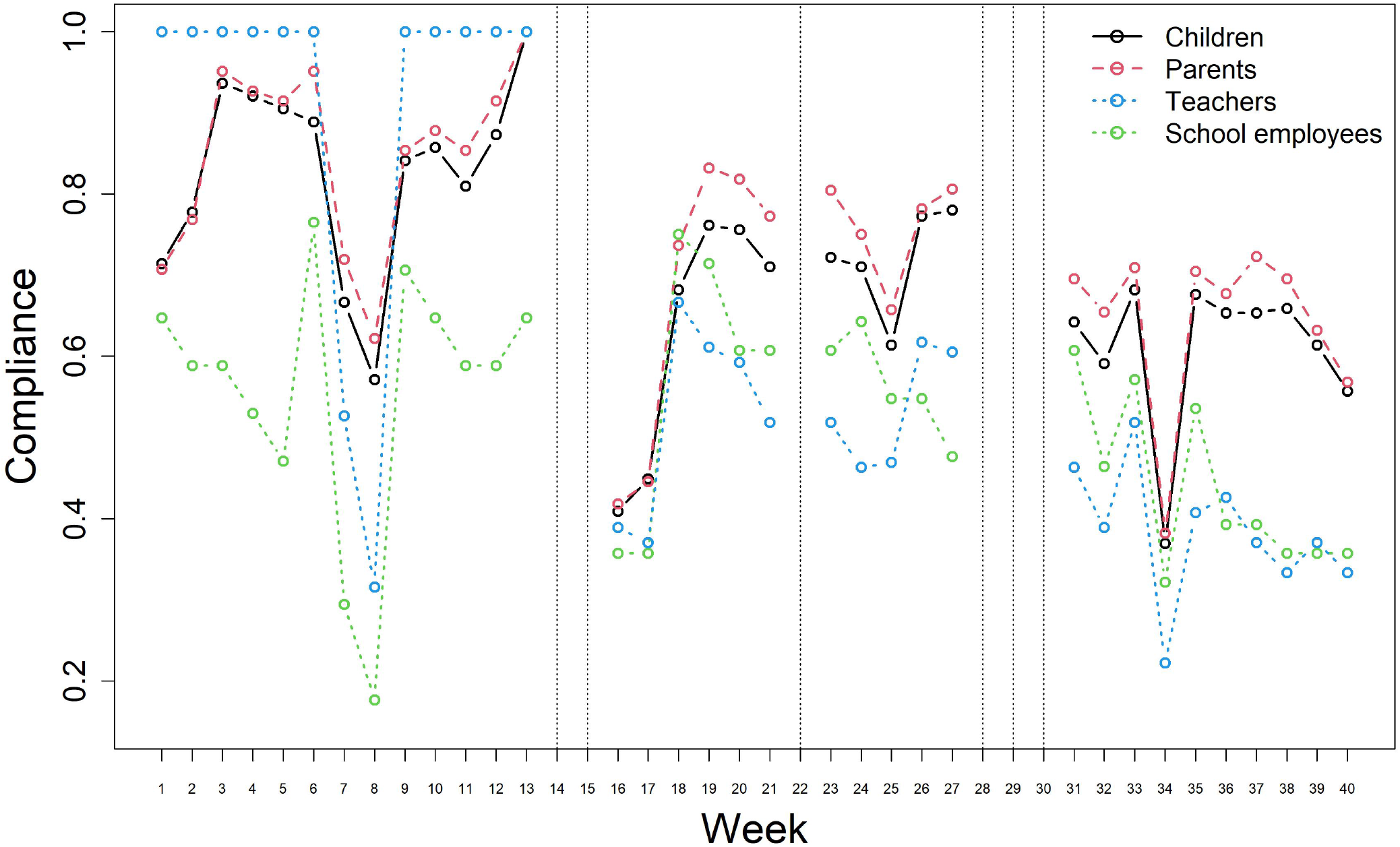
Compliance to the study protocol over time by participant group. Compliance is defined as the number of samples obtained divided by the expected number of samples for each week. Dotted vertical lines indicate weeks of school holidays when no samples were collected.

### Sequence alignment and phylogenetic analysis

Analysis of the whole genome sequences (WGS) was performed as described in Methods. Forty WGS were available for 36 distinct cases (59% of all cases). Quality assessment of the raw sequence data identified no issues concerning frameshifts, premature stop codons, mixed sites, clusters of mutations, or excess private mutations. There were four individuals from whom two samples were sequenced. In the following analyses, the earliest sampled WGS for each confirmed case was used. The reasons for excluding the later sequenced samples were (i) identical sequence, (ii) only a partial genome due to high Ct sample, and (iii) a high proportion of ambiguous nucleotides. For all sequences, the region encoding the envelope protein was excluded due to alignment issues caused by missing nucleotides in 12 sequences. Mutations in the envelope protein have only been reported in the Beta (B.1.351) variant [13], which has not been observed in this study. The most frequently observed amino acid changes compared to the reference sequence were the P4715L mutation in the ORF1ab gene occurring in 100% of the sequences, and the D614G mutation in the spike glycoprotein occurring in 91.7% of the sequences (Supplementary Fig. S4). A previous study has found these to be the most prevalent mutations in WGS sampled between December 2019 and September 2020 (i.e., wild type SARS-CoV-2) [14]. The ORF8 Q27* knockout mutation was detected in the 7 sequences from the March cluster and is a known defining mutation for the Alpha VoC [15]. The best fitting substitution and rate heterogeneity across sites (RHAS) model was a TN93 model with empirical base frequencies and no rate heterogeneity across sites [16]. The sequenced cases belong to four different pangolin lineages, i.e., B.1.221 (47.2%), B.1.160.28 (30.6%), B.1.389 (2.5%), and B.1.1.7 (19.4%), as shown in the time-scaled phylogenetic tree (Figure 2).

**Figure 2:**
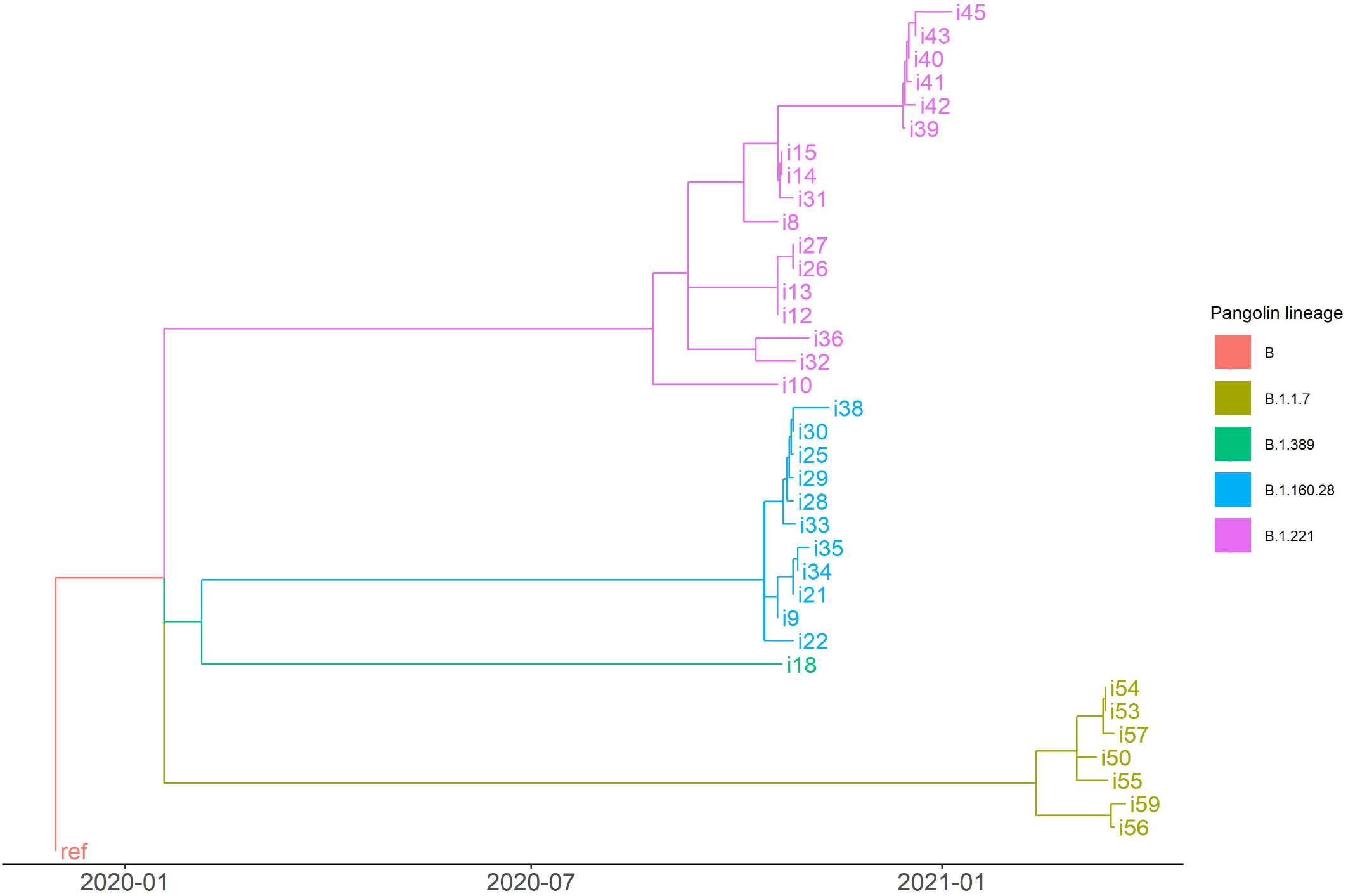
Time-scaled maximum likelihood tree. The tree is rooted on the reference sequence (ref) which represents an isolate obtained early in the pandemic. Branches are colored according to pangolin lineage.

### Outbreak reconstruction

To gain insight on the extent of transmission within the school, reconstruction of the outbreaks was performed using two different models, Outbreaker2 and SCOTTI (see Methods for a description of how both models were implemented) [17–19]. Trace plots indicated adequate convergence of the MCMC chain for both models (Supplementary Figs. S5 and S6). Supplementary Table S1 shows the assumed prior distributions and posterior estimates with 95% credible intervals (CrI) for the parameters of the Outbreaker2 model under a baseline and sensitivity scenario. In the consensus transmission tree, defined as the tree with the modal posterior infector for each case, 27 direct transmission events occurred in school, 14 in households, and 15 were indirect transmission events. Of the direct transmission events, 12 (29.3%) occurred among children, 16 (39.0%) among adults, 10 (24.4%) from child to adult, and 3 (7.3%) from adult to child (Figure 3). Based on the posterior distribution of ancestors, direct transmission was most likely to have occurred between children or from children to adults during the March outbreak when the Alpha VoC was circulating (Supplementary Fig. S7). In October (case 1 to 38) and December (case 39 to 47) it was more likely to have unobserved intermediate cases (i.e., *κ >* 1) compared to March (case 48 to 59) when sampling was done three times a week (Supplementary Fig. S8). The use of uninformative priors did not impact estimates of the model parameters (Supplementary Table S1). Another sensitivity analysis assuming the generation and time-to-collection interval to be uniformly distributed over 10 days led to similar results, with a slightly higher estimate for the proportion of cases sampled.

**Figure 3:**
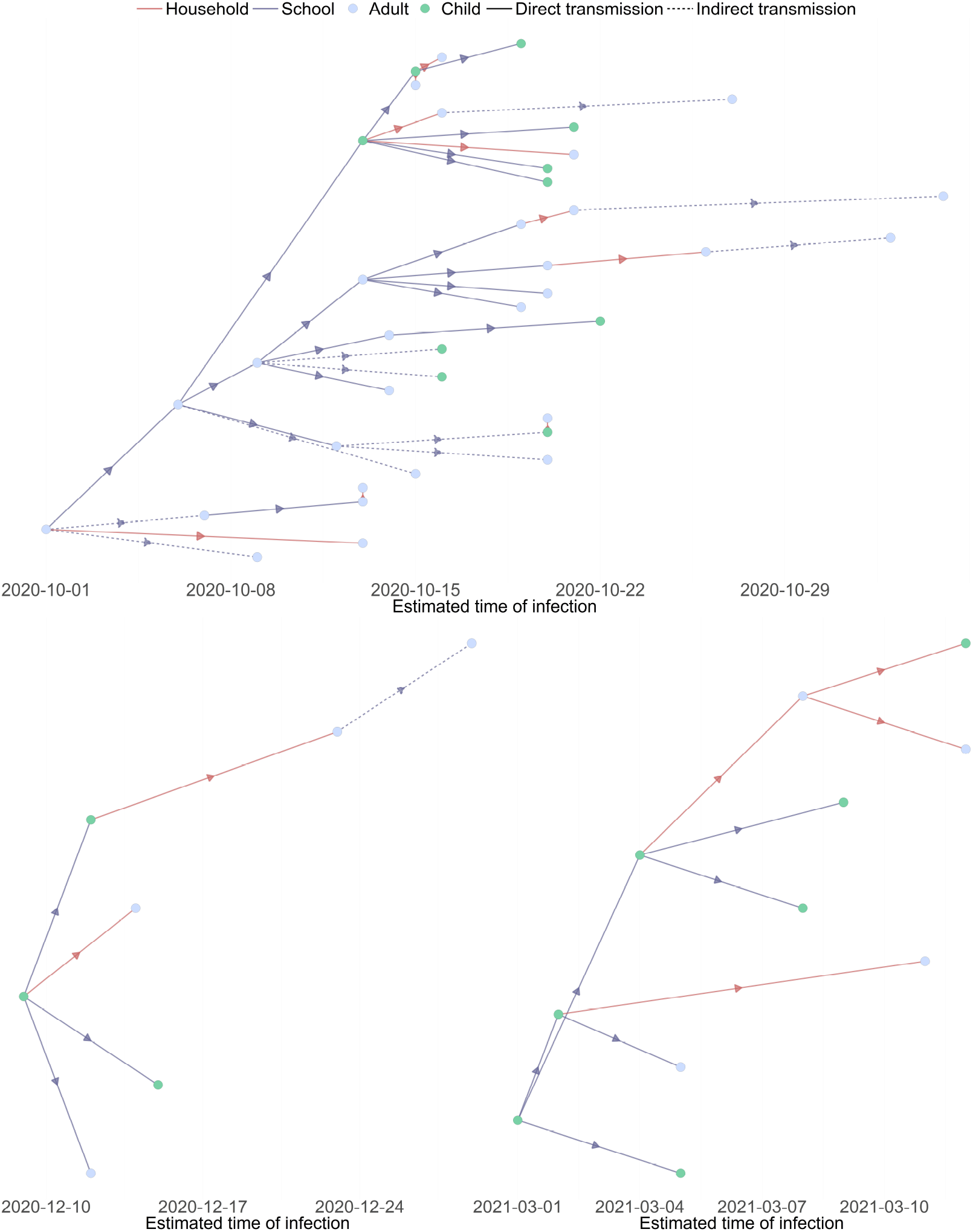
Consensus transmission tree under the baseline scenario from the Outbreaker2 model. The consensus tree is defined as the tree with the modal posterior ancestor for each case. Indirect transmission (dashed line) means at least one unobserved intermediate case is present.

Supplementary Table S2 shows the posterior estimates and 95% highest posterior density (HPD) intervals for the parameters of the SCOTTI model for each of three separately analysed time periods. When using SCOTTI, for 24 (66.7%) of the 36 cases included in this analysis, the highest probability of direct infection was linked to transmission in school. Especially for the cases linked to the October cluster, SCOTTI assigned only very low probability to observed infectors, with most cases assigned to have been infected by a non-sampled case. Because SCOTTI only uses the cases for which a genome sequence is available, these non-sampled cases could in fact be the observed cases for which no sequence was available and that were assigned as maximum posterior ancestry by Outbreaker2.

Uncertainty related to inferred ancestry was quantified using the Shannon entropy of the posterior distribution of potential infectors for each case [20], with a higher entropy indicating more uncertainty in ancestry assignment (see Methods) [17]. For the Outbreaker2 model, the average entropy among all cases was 1.43 when including and 1.60 without including sequence data. For SCOTTI, the average entropy was 1.46 among the 36 cases included in this analysis. Using Outbreaker2 without sequence data, most uncertainty is observed for the cases in October (case 1 to 38), and this uncertainty is in general slightly reduced when including sequence data in the outbreak reconstruction. For most cases observed during the December and March outbreaks, there is more uncertainty in ancestry assignment when using SCOTTI compared to Outbreaker2 including sequence data (Figure 4).

**Figure 4:**
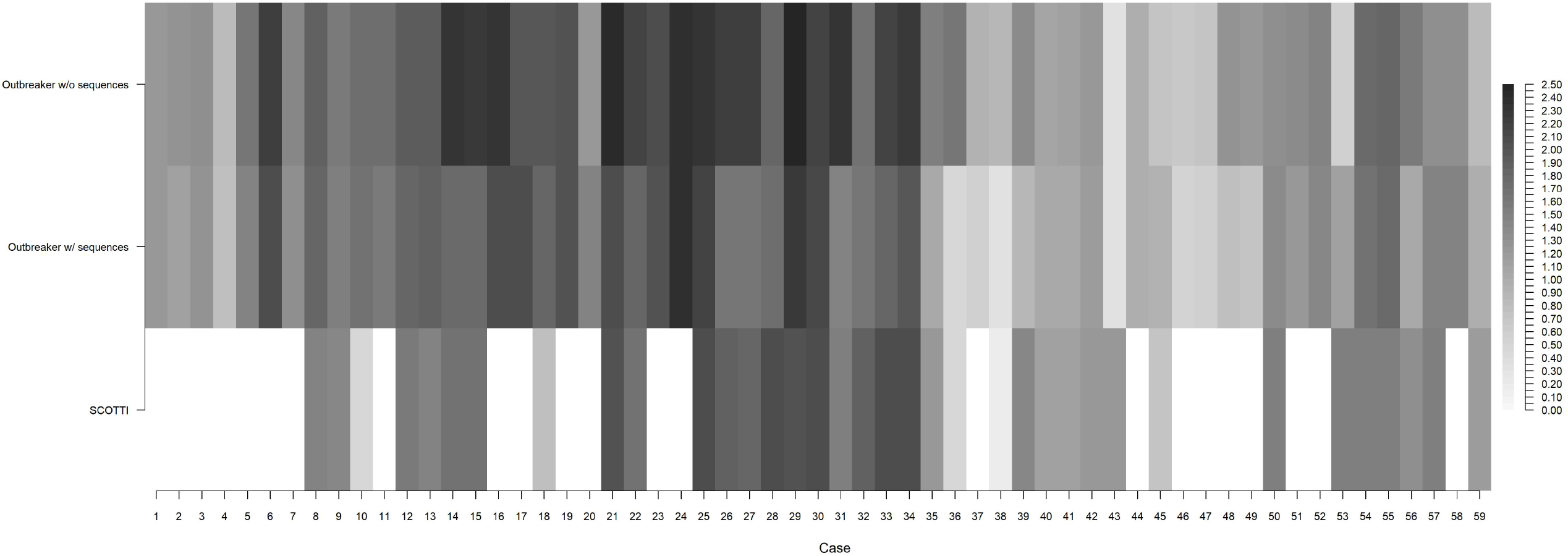
Uncertainty in ancestry assignment for the different models. For each case, uncertainty is quantified as the Shannon entropy of the posterior distribution of potential infectors. Only 36 cases are included in the SCOTTI model. Higher values indicate a larger number of plausible infectors, hence higher uncertainty in ancestry assignment.

### Simulation study

A previously developed individual-based model representing a population of primary school children and their teachers was used to investigate how the accuracy of estimated weekly positivity rates depends on the proportion of a school that is sampled in a repetitive screening strategy (see Methods for a description of the model) [9, 21]. The probability of symptomatic infection was set to 65% for children and 85% for adults, as observed in the surveillance study. We assumed that 10% of children acquired immunity from previous infection, and that 30% of adults were immune due to previous infection or vaccination. We found that the true positivity rate is underestimated during the first weeks of an outbreak. After some introductions, the deviation between true and observed positivity rates as well as the variation in this deviation decreases for an increasing sampling proportion. Repetitive testing at a frequency of twice instead of once weekly generally does not result in a big improvement in estimation of the positivity rate. In contrast, a strategy of only testing and isolating symptomatic individuals results in a consistent underestimation of the true positivity rate (Figure 5). Lowering the probability of symptomatic infection (to 30% for children and 50% for adults) or increasing the proportion of immune adults to 50% did not have an impact on these results. Estimation of the weekly positivity rate in children was in general more accurate than the rate in adults (Supplementary Fig. S9).

**Figure 5:**
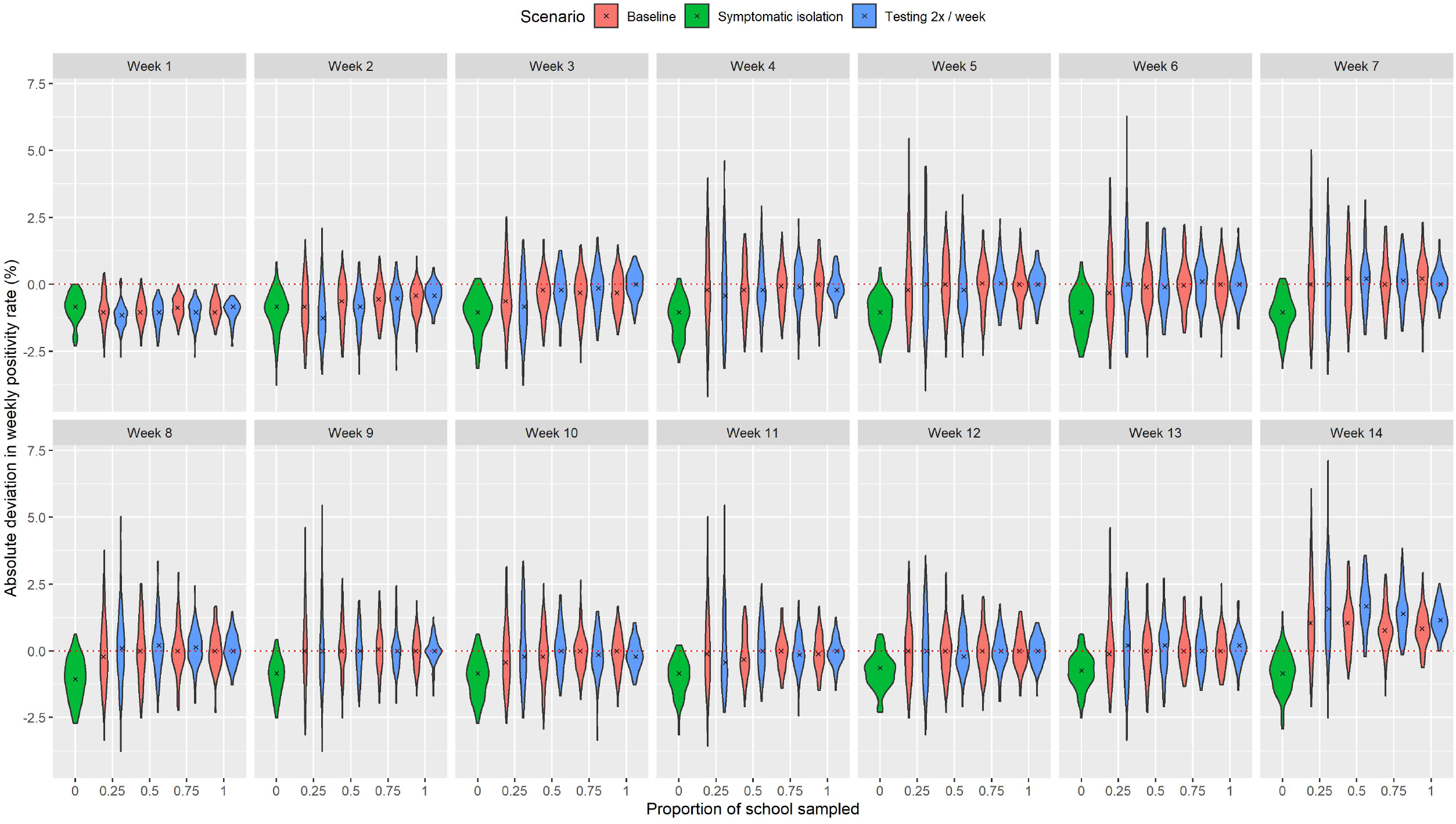
Accuracy of estimated weekly positivity rates. Accuracy is defined as the absolute deviation (observed minus true positivity rate) in estimates of the weekly positivity rate (in %) for different proportions of the school sampled. Black crosses represent the median deviation among 100 simulated outbreaks. In the baseline scenario, testing was performed once a week.

## Discussion

In the present study SARS-CoV-2 infection was equally present among adults and children, consistent with findings from previous studies [7, 10, 22]. We found no significant difference in the proportion of asymptomatic cases between children and adults during the second part of the study, while it was observed that children were more often asymptomatic during September – December 2020 [10]. This may be explained by circulation of the Alpha VoC in the cluster observed in March 2021, for which it has been shown that children have a higher probability of developing symptoms compared to wild-type SARS-CoV-2 [23]. In line with a previous study that found no effect of age on Ct values after adjusting for the effects of self-reported symptom status and number of positive genes [24], we observed no difference in Ct values between children and adults. As a proxy for viral load, Ct values may inform an individual’s infectiousness [25], suggesting that children and adults may have been equally infectious in this study. From April 2021 onward, only three sporadic cases were observed. Vaccination of adults could have had a protective effect on the school community. As a lot of parents included in this study are healthcare workers, about half of them had been vaccinated relatively early on. However, only 14.1% of the vaccinated adults were teachers, and contact between parents and children that are not their own was assumed to have been limited during the study period. Even if all teachers would have been vaccinated, they only make up a small part of the school population and have lower contact rates with students compared to students amongst each other, hence vaccination of children may be needed to limit infections in schools and decrease the risk of importation at the community level [8].

Because the two models used for outbreak reconstruction differ in their estimation procedure they can only be compared in a broad sense in terms of inference of the transmission network [26]. The main purpose of outbreak reconstruction in this study was to infer the extent of transmission within the school environment. Using SCOTTI, we found that for 66.7% of cases transmission most likely occurred at school. Similarly, using Outbreaker2 we estimated that 65.9% of direct infection occurred within the school and furthermore that more than half of direct transmission events most likely originated from a child. These results imply that children, as well as teachers, pose a risk of bringing infection into their households, from which it can spread further into the community. In line with this, a recent retrospective cohort study in students, school personnel, and their household members found that younger age groups had a high contribution to the spread of infection [27]. Similarly, two recent studies in The Netherlands and Switzerland found evidence of large-scale transmission among children, school personnel, and introduction into their households during dominance of the Alpha VoC [23, 28]. Uncertainty in outbreak reconstruction was lowest for the Outbreaker2 model that uses genomic sequence data in addition to epidemiological data. However, the average entropy for SCOTTI, which does not use information on contacts, and Outbreaker2 was similar, suggesting that the addition of contact data may not be very informative in a school setting.

From January to June 2021, we observed lower positivity rates compared to before January 2021, reflecting the lower background incidence in the Belgian population at that time. In addition, positivity rates were higher for children compared to adults from January to June 2021. In our simulation study, observed weekly positivity rates were found to be a good approximation to the true weekly positivity rate, especially in children, even when only a small proportion of the school is tested regularly. In practice, this can be used to evaluate and if necessary adapt mitigation measures in school. Symptomatic isolation has already been shown not to be very effective in reducing SARS-CoV-2 infections in schools [8, 9], and we found that data obtained from such a strategy results in consistent underestimation of positivity rates. Further research is needed to investigate how the accuracy of outbreak reconstruction depends on which proportion of a school population is sampled and how many of these samples are sequenced.

Adherence to the screening protocol has been assumed crucial for a repetitive testing strategy to be effective [8]. We observed moderate-to-high adherence to either once or twice a week testing, with adherence in general being higher among children and parents compared to school staff. In contrast there was also substantial loss to follow-up. Although this cannot be directly inferred from the available data, possible reasons for dropout include fatigue, vaccination, stress generated by positive results, and previous infection of the individual or in their household leading to a sense of protection. A recent modeling study found that regardless of the level of compliance, repetitive testing always leads to a greater reduction in final outbreak size compared to reactive screening, although the reduction in final size increased with higher levels of compliance [9]. When social mixing between classes is high, as in primary schools, other classes may have already been affected before the first case is detected, making a reactive screening strategy less efficient [29]. In contrast, with repetitive testing, more cases that would otherwise go unnoticed will be detected and isolation can be applied only to those cases during their infectious period, instead of quarantining the entire class.

This study has several limitations. Assessment of symptoms was retrospective and hence could be biased by the knowledge of having been infected. Especially in children, it may have been the case that very mild symptoms with a duration of less than one day have been reported. The Outbreaker2 model makes a number of simplifying assumptions [17]. Contacts are undated, hence the model does not consider that these contacts only could have led to transmission if they occurred during the infectious period of the infector. However, in a school setting, the same individuals will be in contact with each other almost daily. Possible within-host evolution is not accounted for, hence the genetic likelihood only depends on the number of transmission events separating two cases and not on time. In addition, the genetic likelihoods are assumed to be independent while in fact the genetic relatedness between two cases, A infecting B, is dependent on the infector of case A. SCOTTI overcomes some of these limitations by explicitly using the sequence data rather than only the pairwise genetic distances and allowing transmission to occur only within a specified exposure window. However, the assumption that transmission is equally likely between every pair of cases during their exposure window may be reasonable for transmission among children in the school environment but not for transmission between children and adults who are not their teacher or parent. Furthermore, SCOTTI does not include the non-sequenced cases, thereby excluding a large part of the available information which is reflected in the high probabilities assigned to non-sampled cases being the most likely ancestor.

Despite these limitations, the present study shows that it is worthwile to implement repetitive screening in a school setting. In addition to reducing infections, important insights can be obtained on the role of schools in transmission of infectious diseases such as the extent of transmission happening within the school environment. In particular, we found that the majority of SARS-CoV-2 transmission occurred in school, with importation to students’ and staff members’ households. Challenges to implementing such a strategy include the cost of PCR testing as well as quickly obtaining test results to ensure fast isolation of positive individuals in order to prevent onward transmission within as well as outside the school. Testing capacity can be quickly overwhelmed when case numbers are high. In periods of low incidence, pooled testing could be a more cost-effective approach [30]. Alternatively, the use of antigen tests could be considered [29]. Besides educational disadvantages, it is important to minimize SARS-CoV-2 infections in children in order to avoid possible long-term health disadvantages caused by conditions such as long-COVID or multisystem inflammatory syndrome, which can occur even after mild or asymptomatic disease [6, 31, 32].

## Methods

### Sample collection

Prospective surveillance of SARS-CoV-2 throat carriage was performed among children, their parents, and employees in a single primary school in Liège (Belgium) from September 2020 to June 2021. Weekly screening for SARS-CoV-2 infection through throat washing was performed from September to December 2020. From January to June 2021, screening was done twice a week using the same protocol. An outbreak in March 2021 prompted a change in strategy to three samples a week for two consecutive weeks. No screening was performed during school holidays. All adults provided informed consent, also on behalf of their children. Mitigation measures evolved during the course of the study, following national guidelines for testing and quarantine in primary schools (Supplementary Table S3). When tested positive for SARS-CoV-2, participants were called by the investigators to fill in a questionnaire about the timing of symptom onset and symptom duration. In addition they were instructed to isolate in order to limit further spread of the virus. Family members were quarantined according to national guidelines and advice was given on how to try preventing transmission within the household. For full details on the study design, we refer to Meuris et al. [10].

### SARS-CoV-2 detection and sequencing

Quantitative reverse transcription-PCR was used to detect SARS-CoV-2 and sequencing was performed as described by Freed et al. [33]. RNA was extracted from throat washing (300*μl*) via a Maxwell 48 device using the Maxwell RSC Viral TNA kit (Promega) with a viral inactivation step using Proteinase K, following the manufacturer’s instructions. RNA was eluted in 50*μl* of RNAse free water. 3.3*μl* of the eluted RNA was combined with 1.2*μl* of SuperScript IV VILOTM Master Mix and 1.5*μl* of H2O to carry out Reverse Transcription. This was incubated at 25°C for 10 minutes, 50°C for 10 minutes, and 85°C for 5 minutes. PCR was carried out using Q5^®^ High-Fidelity DNA Polymerase (NEB) and primers to obtain 1200bp amplicons as described by Freed et al. [33]. PCR conditions followed the recommendations in the sequencing protocol of the ARTIC Network. The samples were multiplexed following the manufacturer’s recommendations using the Oxford Nanopore Native Barcoding Expansion kits 1-12, 13-24, and 96, in conjunction with Ligation Sequencing Kit 109 (Oxford Nanopore). Sequencing was carried out on a Minion using R9.4.1 flow cells.

### Statistical analysis

SARS-CoV-2 positivity in adults and children was compared using a mixed effects logistic regression model with random intercepts for classroom and household, accounting for the clustering of individuals. For all other analyses, differences between two groups were compared using Pearson’s chi squared, Fisher’s exact, Student’s t, or Mann-Whitney U-tests, as appropriate. Results were considered statistically significant at *p* < .05. Analyses were performed using R version 4.1.0.

### Sequence alignment and phylogenetic analysis

Quality assessment of the whole genome sequences was performed using Nextclade [34]. The sequences were then aligned to the Wuhan-Hu-1 (NC_045512.2) reference sequence using VIRULIGN, which computes a codon correct multiple-sequence alignment relative to this reference sequence [35, 36]. The maximum likelihood phylogenetic tree was inferred using IQ-TREE v1.6.12 [37] with ultrafast bootstrap approximation (UFBoot) using 1000 bootstrap replicates to assess branch support [38]. ModelFinder [39] was used to find the best fitting substitution and rate heterogeneity across sites (RHAS) model based on a full tree search for each combination of models. The Bayesian Information Criterion (BIC) was used to identify the optimal combination. TreeTime v0.8.5 [40] was then used to create a time-scaled phylogeny based on the maximum likelihood tree, rooted on the reference sequence which represents an isolate obtained early in the pandemic.

### Outbreak reconstruction

Reconstruction of the school outbreaks was done to gain insight on the extent of transmission within the school environment. We used the Outbreaker2 model, which combines information on the generation interval and contact patterns with a model of sequence evolution in a Bayesian framework [17, 41]. The model considers, for each case *i* = 1, …, *N*, the probability of a proposed transmission history given the sample collection time *t*_*i*_ and a genome sequence *s*_*i*_. A contact model accounting for partial sampling and the presence of non-infectious contacts between cases is also included. Transmission events are modeled using augmented data, i.e., a case’s infection time 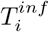, most recently sampled ancestor (MRSA) *α*_*i*_, and the number of generations (*κ*_*i*_ *≥* 1) separating *i* and *α*_*i*_. We assumed the generation interval to be Gamma-distributed with a mean of 4.7 days and standard deviation of 2.5 days [10]. The distribution of the time-to-collection was assumed to follow a Gamma distribution with a mean of 7 days and standard deviation of 2 days. In a baseline scenario, prior distributions supporting higher values for the proportion of contacts reported (*ϵ*) and lower values for the probability of non-infectious contacts (*λ*) were chosen in order to emphasize reported contacts when assigning ancestries (Supplementary Table S1) [17]. For the mutation rate an exponential prior with a mean of 10^*−*5^ substitutions site^*−*1^ generation^*−*1^ was used [42]. The MCMC chain was run for 10^6^ iterations with a thinning frequency of 1/100 and a burn-in of 10%. A sensitivity analysis assessing the impact of the choice of prior distributions was performed using a flat Beta(1,1) prior for each of the parameters *π, ϵ*, and *λ*.

For comparison to the Outbreaker2 model we used SCOTTI which, as part of the BEAST2 platform for Bayesian phylogenetic analysis, is based on a structured coalescent model that enables outbreak reconstruction while accounting for within-host evolution and nonsampled cases [18, 19, 43]. Each case is modelled as a distinct population, and transmissions between cases are modelled as migration events. Non-sampled cases are modelled by dynamically increasing or decreasing the number of populations. The model assumes that transmission is a priori equally likely between every pair of cases, i.e., the migration rate is the same between every pair of cases for the time they are both exposed. It is further assumed that all cases have the same, constant, within-host pathogen evolution dynamics. In addition to genome sequences, SCOTTI incorporates epidemiological information by only allowing cases to transmit the virus during a specified exposure window. We defined the exposure window as being from 7 days before symptom onset (or sampling for asymptomatic cases) until two days after the sequenced sample was obtained (i.e., assuming cases self-isolate within two days after the sample was obtained). For cases that had symptom onset more than 5 days after the date of their positive sample, the lower bound of this interval was set to one day before the sequenced sample was taken. The best fitting substitution and RHAS model identified by ModelFinder was used. A strict molecular clock was assumed, using an exponential prior with a mean of 2*×*10^*−*6^ substitutions site^*−*1^ day^*−*1^ [42]. The analysis was done separately for three time periods (i.e., October-November 2020, December 2020, March 2021). To ensure a high enough but realistic upper bound, the maximum number of cases (sampled and non-sampled) was set to 10 times the observed number of sequences, with as minimum the number of sequenced samples during each period. The MCMC chain was run for 2 *×* 10^7^ iterations with a thinning frequency of 1/2000 and a burn-in of 10%.

Convergence of both methods was assessed using trace plots. One case known to have not contributed to transmission and three sporadic cases in May-June not linked to onward transmission within the sampled population were excluded from the reconstruction. Reinfections were considered to be separate cases. For both models, we quantified the uncertainty related to inferred ancestry using the Shannon entropy of the posterior distribution of potential infectors for each case [20]. Given that a case is assigned *K* possible infectors with frequency *f*_*k*_, (*k* = 1, …, *K*), the entropy is defined as

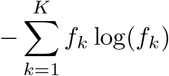

such that an entropy of 0 indicates complete posterior support for a given infector. Higher values indicate a larger number of plausible infectors, hence higher uncertainty in ancestry assignment [17]. In addition we reconstructed the outbreak without using the genome sequence data to investigate whether inclusion of these data reduced the uncertainty in inferred ancestry.

### Simulation study

Full details of the simulation model are available elsewhere [9, 21]. Briefly, children are assigned to a class and interaction among children can occur within as well as between classes (at a level of 20% compared to pre-pandemic behavior). Each class is assigned a teacher, who is assumed to only interact with the children in that class. Symptomatic individuals are assumed to develop symptoms at the peak of their infectiousness, at which time they can be detected and placed in isolation. It is assumed that asymptomatic individuals are as infectious as symptomatic individuals, and that children are half as susceptible as adults. Classes were closed when the number of detected cases exceeds a threshold of 4, while schools were assumed to remain open regardless of the total number of cases. The sensitivity of PCR tests on saliva samples was assumed to be 86% [30]. Disease importation was accounted for by seeding two new cases every week. We simulated 100 outbreaks for different sampling scenarios, considering weekly testing applied to 25%, 50%, 75%, and 100% of the school population at random. As a sensitivity analysis, twice weekly testing was considered. We investigated how the accuracy of estimated weekly positivity rates, quantified as the absolute deviation between observed and true positivity rates, depends on the proportion of a school that is sampled. In addition, we investigated the accuracy of estimated positivity rates among children and adults separately.

## Data Availability

Data and computer code used for these analyses will be made available upon publication of the manuscript.

## Acknowledgements

The study was funded by Fondation Léon Fredericq and by the Liège University Hospital Research funds. P.J.K.L. gratefully acknowledges support from the Research Foundation Flanders (FWO) via postdoctoral fellowship 1242021N (P.J.K.L.). P.J.K.L. also acknowledges support from the Research council of the Vrije Universiteit Brussel (OZR-VUB) via grant number OZR3863BOF, and from the Flemish Government through the AI Research Program. G.D. is an FNRS postdoctoral clinical master specialist. P.L. acknowledges support by the European Research Council under the European Union’s Horizon 2020 research and innovation programme (grant agreement number 725422 - Reservoir-DOCS) and the Research Foundation – Flanders (‘Fonds voor Wetenschappelijk Onder-zoek – Vlaanderen’, G066215N, G0D5117N and G0B9317N). N.H. and A.T. acknowledge funding from the European Union’s Horizon 2020 research and innovation programme - project EpiPose (Grant agreement number 101003688). This project was supported by the VERDI project (101045989), funded by the European Union. Views and opinions expressed are however those of the author(s) only and do not necessarily reflect those of the European Union or the Health and Digital Executive Agency. Neither the European Union nor the granting authority can be held responsible for them. The computational resources and services used in this work were provided by the VSC (Flemish Supercomputer Center), funded by the Research Foundation Flanders (FWO) and the Flemish Government department EWI.

## Author contributions

N.H., P.J.K.L., C.K., A.T., Ch.M., and G.D. contributed to conceptualization of the study. Ch.M., G.D., C.M., M.P.H., S.B., K.D., M.A., and V.B. contributed to acquisition of the data. C.K., A.T., and P.J.K.L. contributed to the analysis code. C.K. performed the analysis. N.H., Ch.M., G.D., C.K., A.T., P.J.K.L, and P.L. contributed to the interpretation of results. C.K. drafted the manuscript. All co-authors critically reviewed and revised the manuscript.

## Ethical approval

This study was approved by the ethics committee of Liège University Hospital (number: 2020-241), the school directors, and the local authorities.

