## Supplementary Information for "Reconstruction of SARS-CoV-2 outbreaks in a primary school using epidemiological and genomic data"

| Scenario | Parameter | Initial value | Prior | Posterior estimate (95% CrI) |
|---|---|---|---|---|
| Baseline | Mutation rate (/site/generation), $\mu$ | 0.00001 | Exp(100000) | 0.00002 (0.000001 - 0.0001) |
| | Proportion of cases sampled, $\pi$ | 0.7 | Beta(5, 3) | 0.66 (0.53 - 0.81) |
| | Proportion of contacts reported, $\epsilon$ | 0.8 | Beta(6, 1) | 0.79 (0.59 - 0.97) |
| | Non-infectious contact probability, $\lambda$ | 0.0001 | Beta(1, 20) | 0.07 (0.05 - 0.10) |
| | Prob. of contact between transmission pairs, $\eta$ | 1 | Fixed | - |
| | Prob. of falsely reporting a contact, $\xi$ | 0 | Fixed | - |
| Sensitivity | Mutation rate (/site/generation), $\mu$ | 0.00001 | Exp(100000) | 0.00002 (0.000001 - 0.0001) |
| | Proportion of cases sampled, $\pi$ | 0.7 | Beta(1, 1) | 0.69 (0.54 - 0.85) |
| | Proportion of contacts reported, $\epsilon$ | 0.5 | Beta(1, 1) | 0.70 (0.48 - 0.92) |
| | Non-infectious contact probability, $\lambda$ | 0.1 | Beta(1, 1) | 0.08 (0.05 - 0.12) |
| | Prob. of contact between transmission pairs, $\eta$ | 1 | Fixed | - |
| | Prob. of falsely reporting a contact, $\xi$ | 0 | Fixed | - |

Table S1: Initial values, prior distributions, and posterior estimates (95% credible interval (CrI)) for the parameters of the Outbreaker2 model under the baseline scenario and sensitivity analysis using uninformative priors for $\pi$, $\epsilon$, and $\lambda$.

| Parameter | Posterior estimate (median and 95% HPD) | | |
|---|---|---|---|
| | October 2020 | December 2020 | March 2021 |
| Mutation rate (/site/day), $\mu$ | $1.4\times10^{-6}$ ($1.5\times10^{-8}$ - $4.1\times10^{-6}$) | $5.0\times10^{-7}$ ($7.2\times10^{-11}$ - $2.5\times10^{-6}$) | $2.5\times10^{-6}$ ($3.1\times10^{-9}$ - $7.8\times10^{-6}$) |
| Effective population size, $N_e$ | 1.47 (0.09 - 20.39) | 0.01 ($1.6\times10^{-6}$ - 2.13) | 0.87 (0.002 - 21.10) |
| Infection rate | 0.005 (0.003 - 0.006) | 0.02 (0.01 - 0.03) | 0.01 (0.009 - 0.02) |
| Number of cases | 141 (60 - 228) | 13 (7 - 41) | 18 (8 - 54) |
| tMRCA (dd/mm/yy) | 17/9/19 (5/1/11 - 20/8/20) | 11/12/20 (16/11/20 - 10/12/20) | 15/2/21 (13/5/20 - 6/3/21) |

Table S2: Posterior estimates (95% highest posterior density (HPD) interval) for the parameters of the SCOTTI model for each of the three periods. In October, three different pangolin lineages were sampled and their tMRCA is close to the wild type SARS-CoV-2 tMRCA, which is not expected to be accurately estimated.

| **Implementation date** | **Mitigation measure(s)** |
|---|---|
| 10 August 2020 | Contact with positive case in the classroom considered low-risk<br>If additional positive case(s) within 14 days, entire class quarantined + symptomatic testing |
| 23 September 2020 | 7-14 days quarantine for children with positive household-contact |
| 10 November 2020 | Extracurricular activities suspended |
| 12 January 2021 | Positive household-contact = high-risk → 10-17 days quarantine<br>In case of suspected positive household-contact, child should stay at home until result is known |
| 26 January 2021 | Child seated next to positive case = high-risk → testing and quarantine<br>Class closure after 4 positive cases<br>School closure when $>$ 3 clusters (i.e. $\geq$ 2 cases in class) or 25% of classes has cluster |
| 15 March 2021 | One-day extracurricular activities allowed again |
| 23 March 2021 | Low-risk contacts tested on day 5<br>All children considered high-risk contact if no fixed places in classroom |
| 1 October 2021 | Symptomatic children isolated as soon as possible<br>Positive household-contact = high-risk → quarantine<br>Class closure after 2 positive cases<br>Obligated mask wearing for teachers |
| 31 October 2021 | Extracurricular activities suspended<br>Lunch inside classroom if no distance between classes is possible |
| 26 November 2021 | Class closure after 2 positive cases<br>Obligated mask wearing for children and teachers<br>Recommendation to perform regular antigen test |
| 6 January 2022 | Class closure after 4 positive cases<br>One-day extracurricular activities allowed again |
| 19 February 2022 | No more mask wearing for children |
| 7 March 2022 | No more mask wearing for teachers |

Table S3: Mitigation measures implemented in primary school, 2020 - 2022.

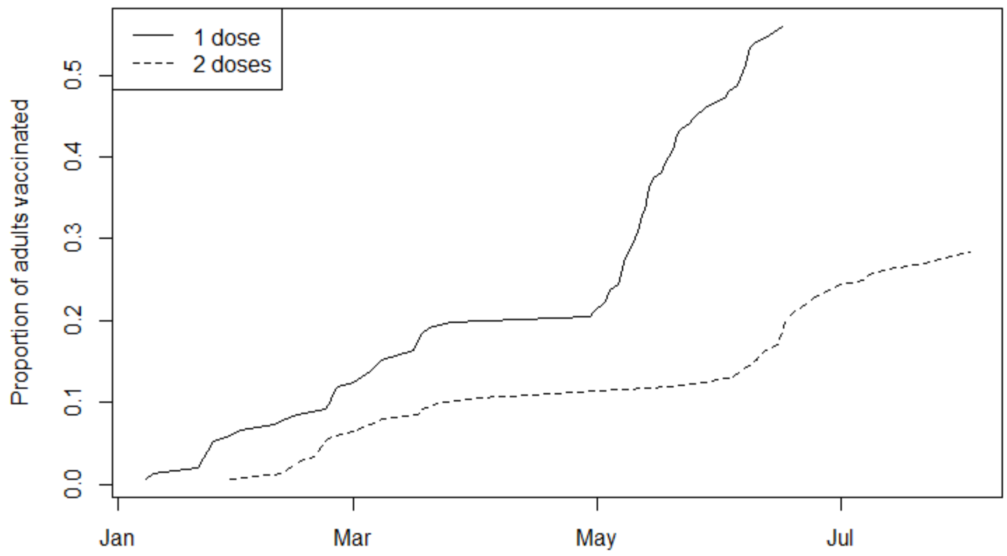


Figure S1: Vaccination coverage among adults in the study population.

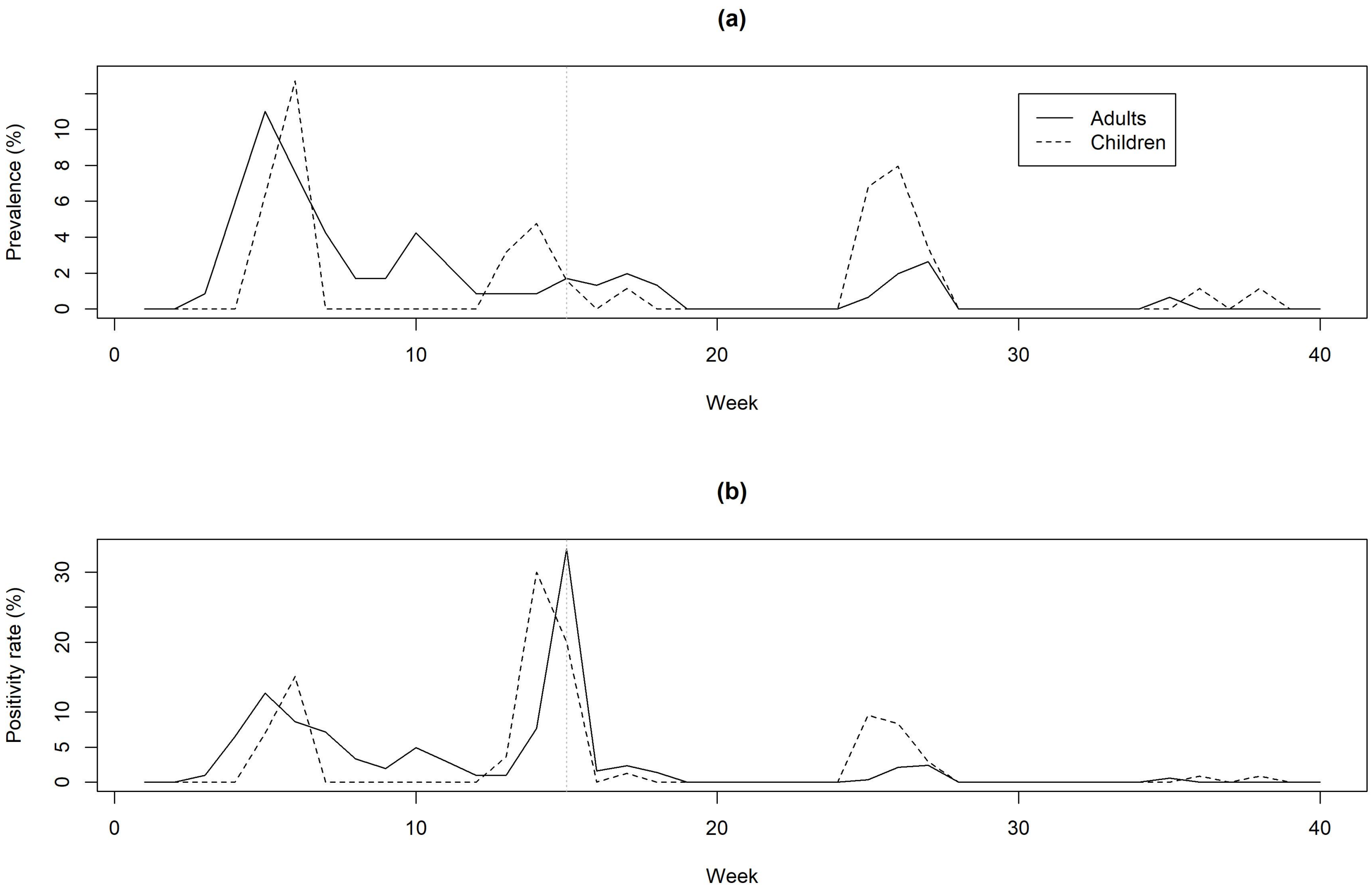


Figure S2: Prevalence (a) and positivity rate (b) over time ($N = 181$ from week 1 to week 15 (i.e., September to December 2020), $N = 240$ from week 16 to week 40 (i.e., January to June 2021)). Sampling was limited to one classroom in weeks 14 and 15. No samples were collected in weeks 22, 28, 29, and 30 due to school holidays.

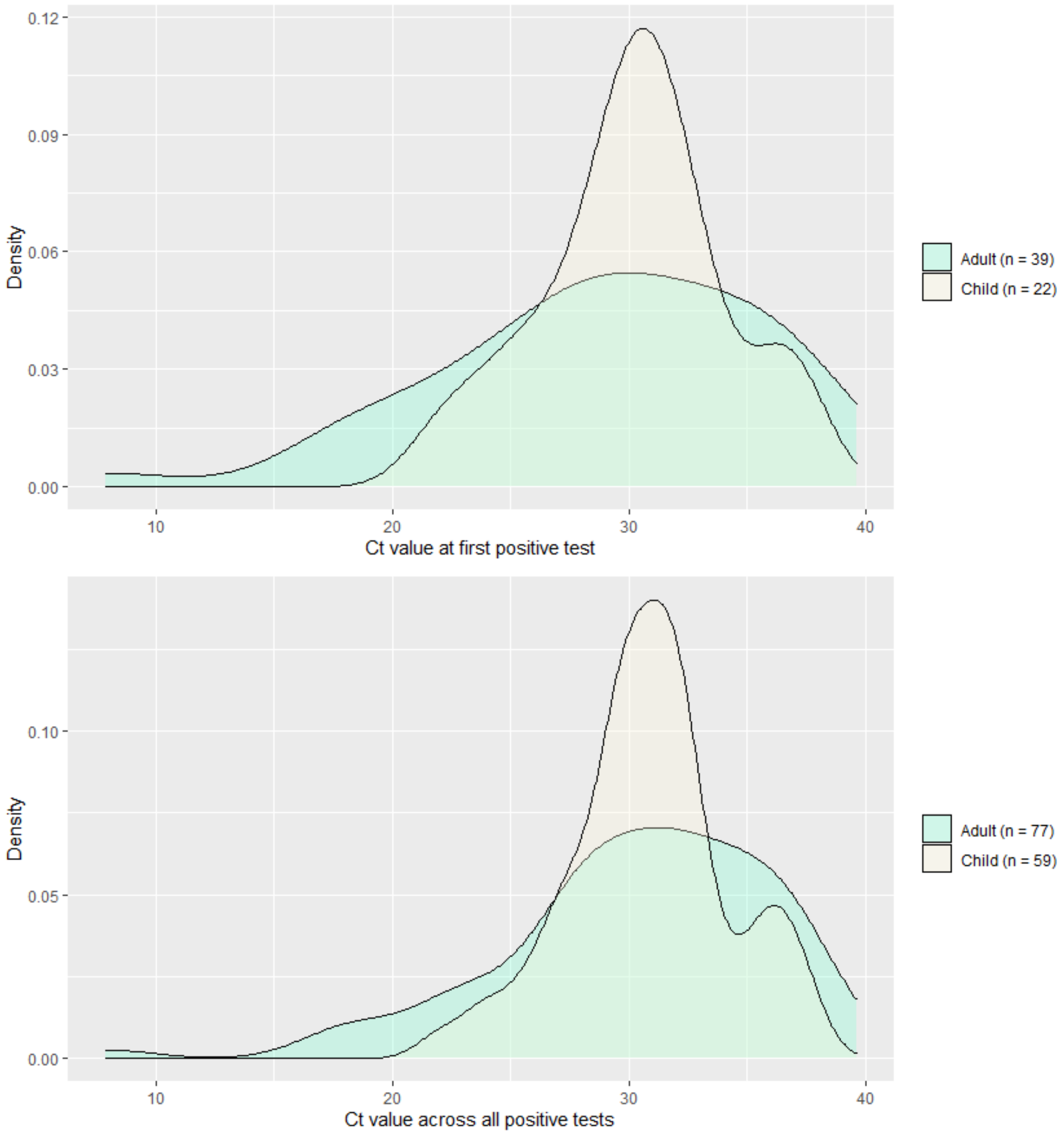


Figure S3: Ct values at first positive test (top) and overall (bottom) for children compared to adults.

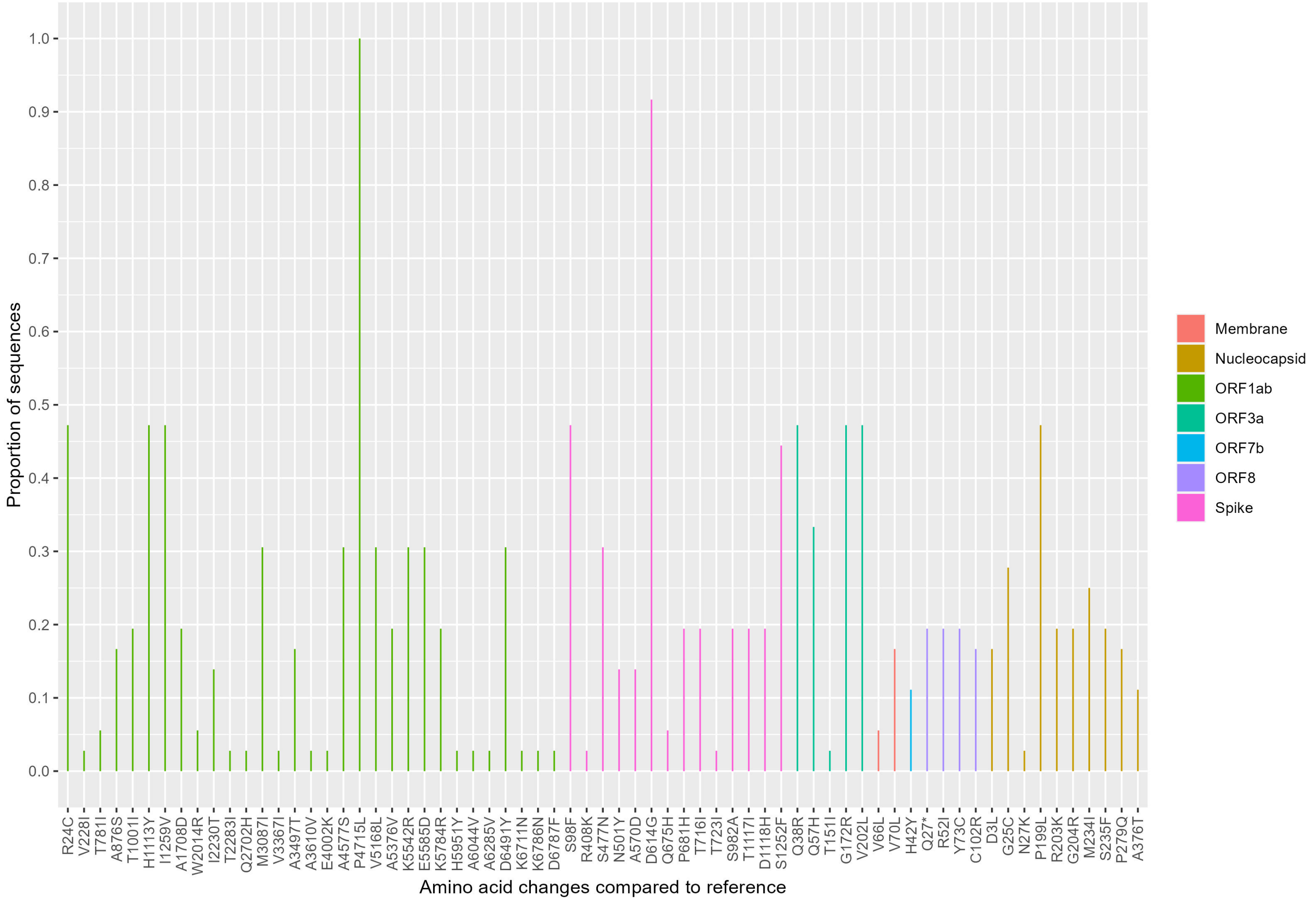


Figure S4: Amino acid changes compared to the Wuhan-Hu-1 reference sequence.

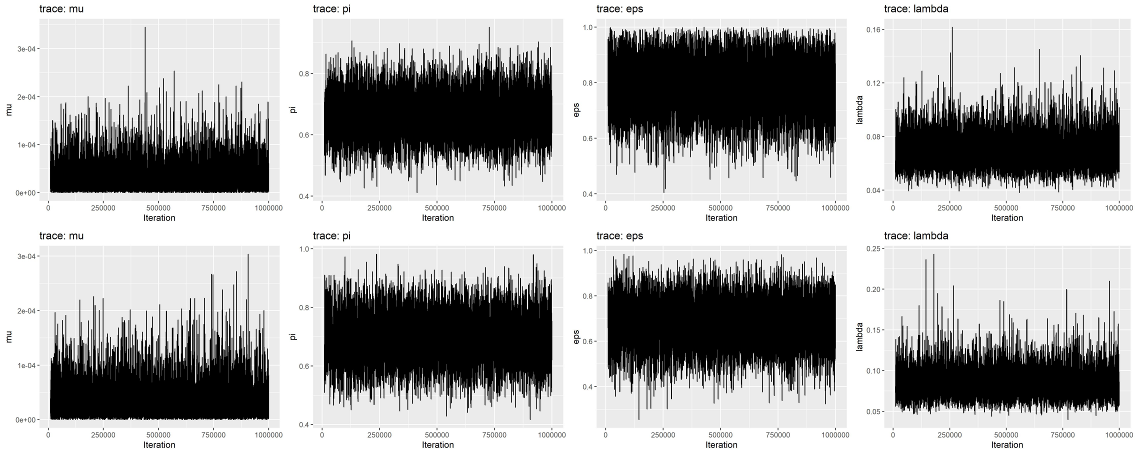


Figure S5: Trace plots showing convergence of the MCMC chain for the baseline (top) and sensitivity (bottom) scenarios of the Outbreaker2 model.

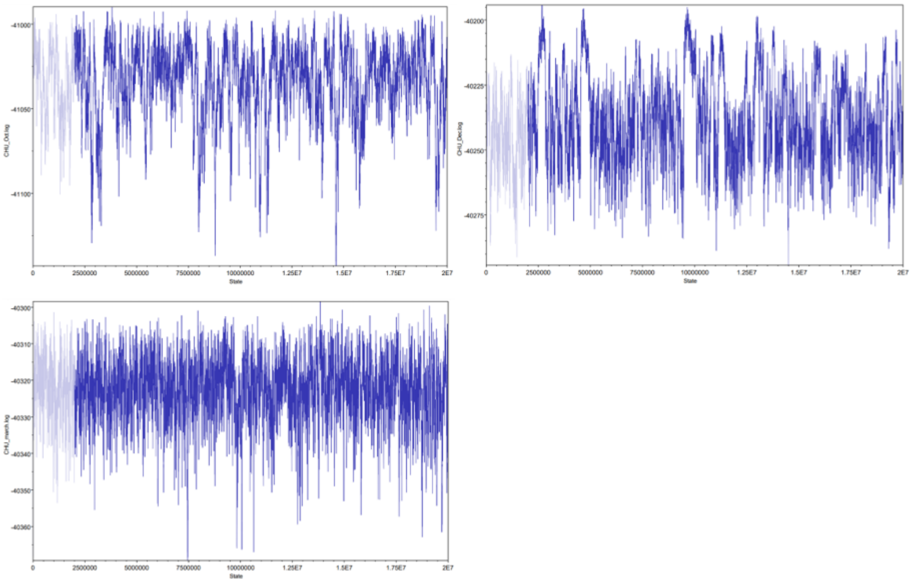


Figure S6: Trace plots showing convergence of the MCMC chain for the SCOTTI model.

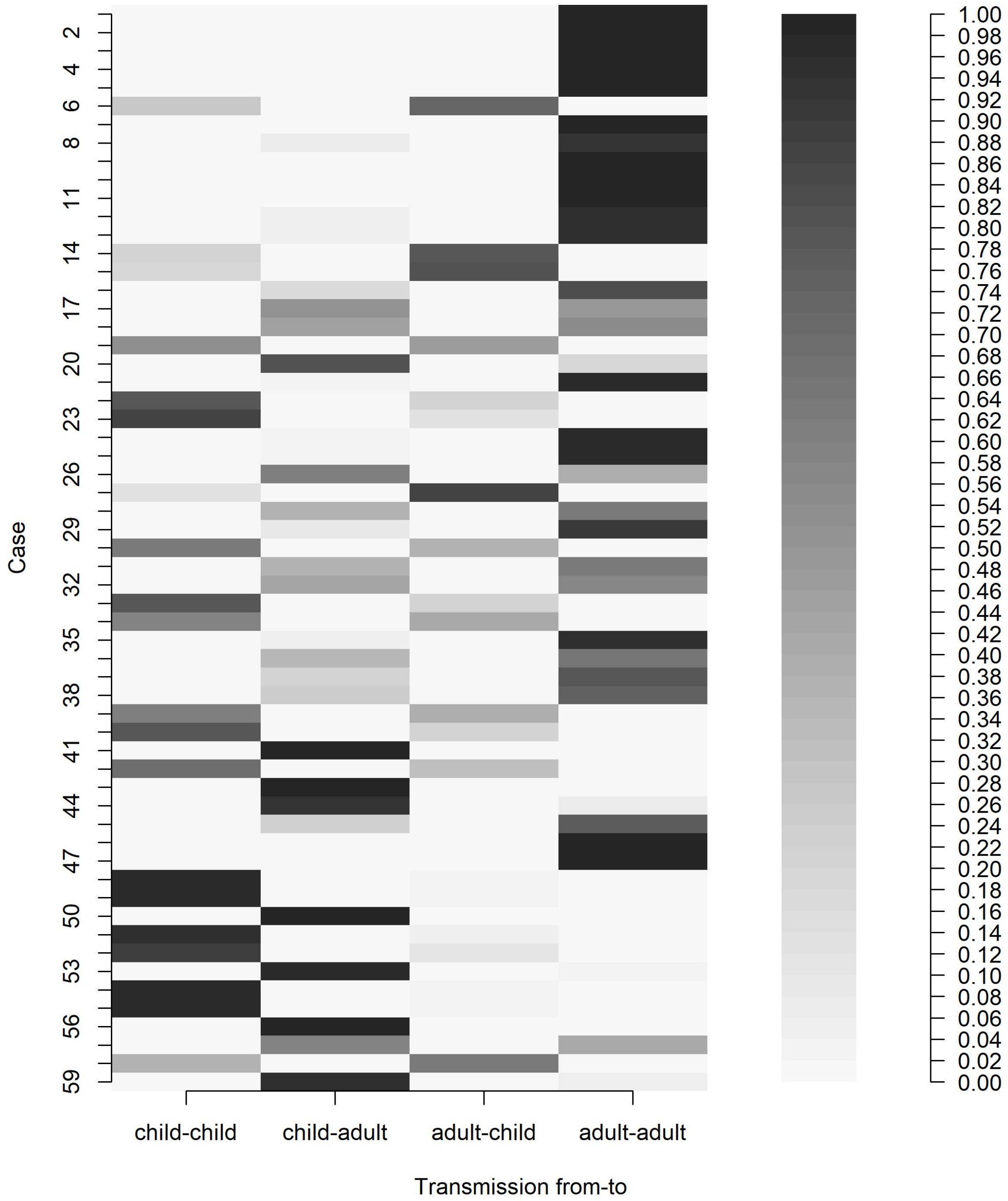


Figure S7: Posterior probabilities of direct transmission events between adults and children under the baseline scenario of the Outbreaker2 model.

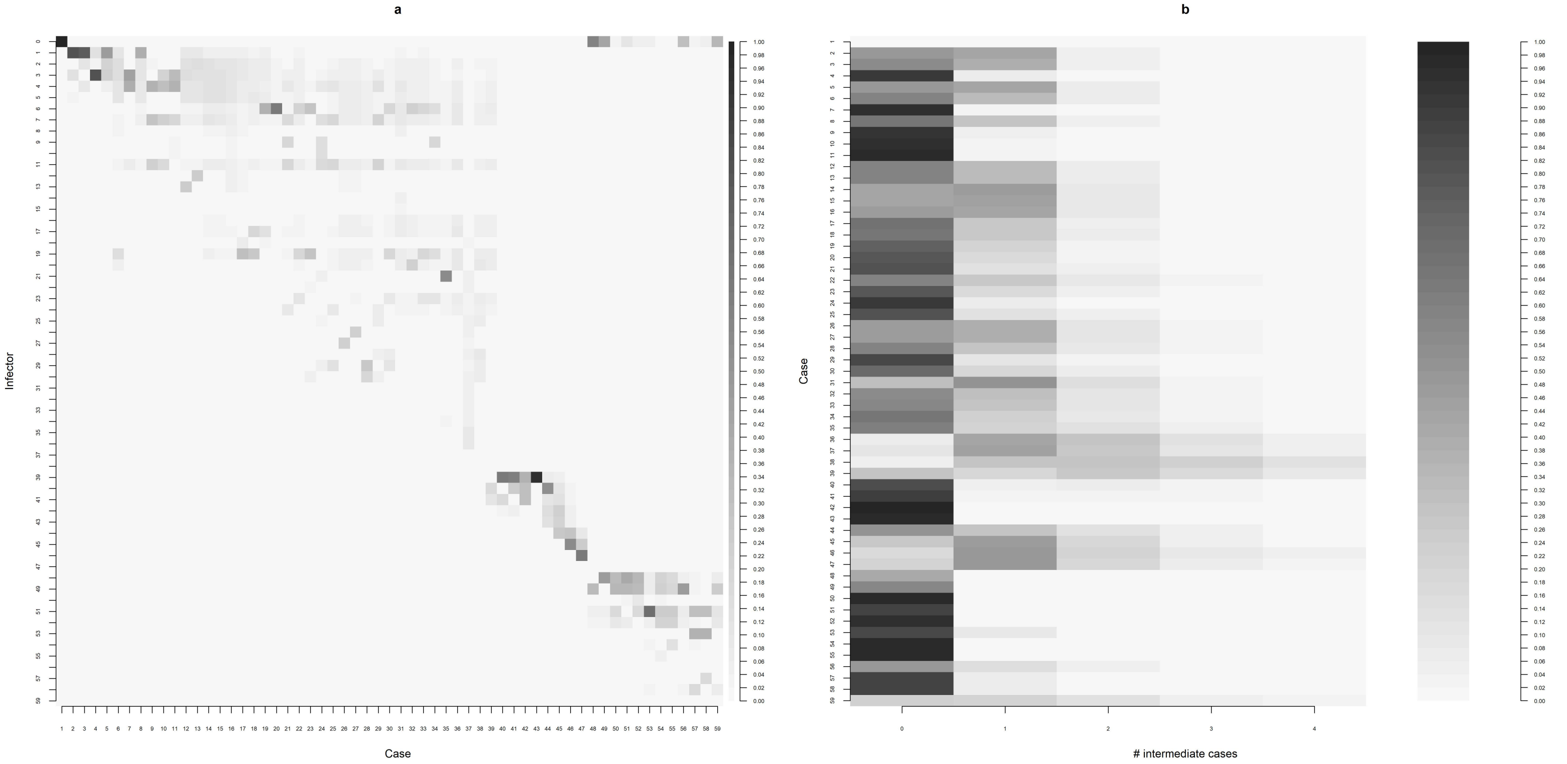


Figure S8: Posterior probabilities for each possible infector (a) and posterior probabilities for the number of intermediate cases (b) under the baseline scenario of the Outbreaker2 model.

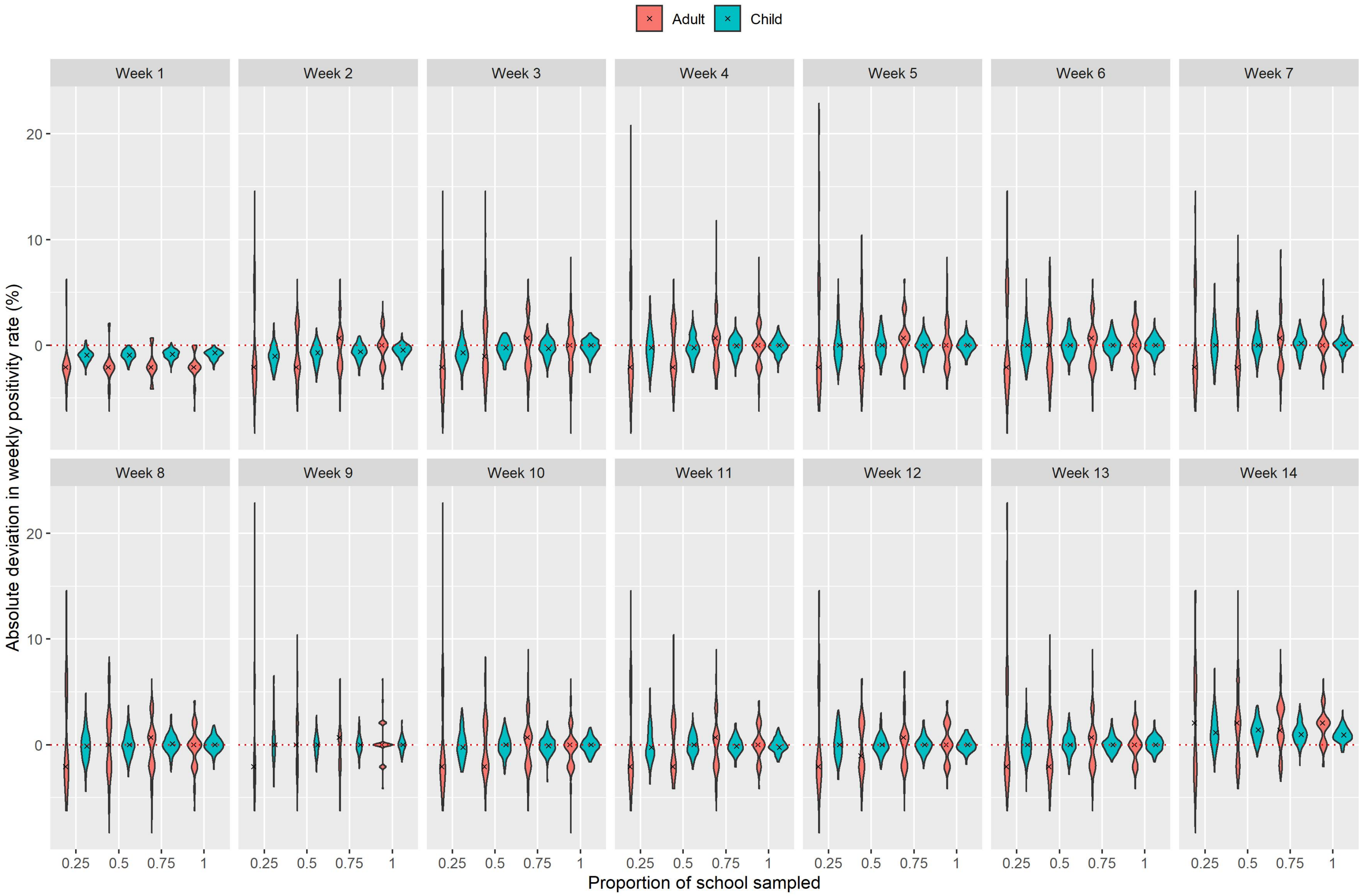


Figure S9: Absolute deviation (observed minus true positivity rate) in estimates of the weekly positivity rate (in %) in adults and children for different proportions of the school sampled under a weekly screening protocol. Black crosses represent the median deviation among 100 simulations.
